# Partnering with Athletes to Assess Risk of COVID-Related Myocarditis

**DOI:** 10.1101/2022.01.29.22270074

**Authors:** Bradley Kay, Attila Feher, Samuel Reinhardt, Jason Cuomo, Stephanie Arlis-Mayor, Matthew Lynch, Kyle Johnson, Phil Kemp, Henry Wagner, Tyler Welsh, Jerome Lamy, Dana Peters, Hamid Mojibian, Lawrence H. Young, Rachel Lampert, Robert McNamara, Lauren A. Baldassarre, Edward J. Miller, Erica S. Spatz

**Author notes:** Corresponding Author: Erica S. Spatz, 789 Howard Ave. Dana 3 Building New Haven, CT 06520. designates co-senior authorship (Lauren A. Baldassarre, Edward J. Miller, Erica S. Spatz). **Funding Source:** Funding provided by Yale New Haven Health System, Department of Radiology and Yale New Haven Health System, Heart and Vascular Center.

## Abstract

**Background:** Myocarditis in athletes is a feared complication of SARS-CoV-2, yet guidelines for screening with cardiac magnetic resonance imaging are lacking. Further, stakeholder involvement in the research is rare.

**Hypothesis:** We sought to determine the rates of cardiac magnetic resonance imaging evidence of SARS-CoV-2 related myocarditis in student athletes. We hypothesized that rates of myocarditis were lower than initially reported and that including athletes on the research team would enhance participant satisfaction and scientific integrity.

**Methods:** Accordingly, when members of a hockey team were infected with SARS-CoV-2, we invited them and their team physicians to be part of the design of a study assessing the incidence of myocarditis. We performed cardiac magnetic resonance imaging on participating hockey players infected with SARS-CoV-2 and compared them to a healthy lacrosse cohort. Participants were given an optional survey to complete at the end of the study to assess their satisfaction with it.

**Results:** Four hockey players and two team physicians joined the study team; eight hockey players and four lacrosse players participated in the study. Zero athletes met imaging criteria for myocarditis; delayed enhancement was observed in seven cases and three controls. Athletes supported sharing the findings with the participants. No athletes reported feeling uncomfortable participating, knowing other athletes participated on the research team.

**Conclusion:** Rates of SARS-CoV-2 myocarditis in young athletes appears to be lower than initially reported. Partnered research is important, especially in populations with more to lose, such as collegiate athletes; future studies should include stakeholders in the study design and execution.

**Key points:** Cardiac MRI findings of myocarditis after COVID infection in young athletes is rare. Subjects of research studies appreciate involvement in the development of the study, and this also builds trust with the research team.

## Background

Myocarditis has been a feared complication of SARS-CoV-2 (COVID-19) infection in competitive athletes as it can increase the risk of arrhythmias, myocardial dysfunction and sudden cardiac death.^1-3^ Initially, the incidence of myocarditis was reported to be between 0.4 and 15%, as based on biomarkers and cardiac magnetic resonance (CMR);^4-7^ yet these studies lacked a standard definition of myocarditis and a control population in some. More recently the focus has been on asymptomatic or mildly symptomatic athletes with COVID-19. In this group, the incidence of abnormal CMR findings appears to be around 2-3%. Still, while there have been no known adverse events in this group, longterm effects are unknown.^4,5,7-11^.

These uncertainties create challenges for screening of myocarditis in asymptomatic or mildly symptomatic athletes, with implications for sports participation and future careers. Specifically, the process for adjudicating what defines myocarditis and thus warrants restriction from sports for 3-6 months - an outcome which can result in deconditioning, decreased psychological well-being, and lost opportunities to compete at both a collegiate and professional level – is not fully known. Additionally, expert recommendations have not included the perspectives of athletes.^12^

Accordingly, when a team of athletes at our institution were contemporaneously infected with COVID-19, we partnered with them to more fully understand the range of findings for myocarditis and to establish guidance for communicating and managing findings.

## Methods

In October 2020, after several members of the men’s hockey team were diagnosed with COVID-19, the Yale Sports Cardiovascular Medicine Team partnered with volunteer members of the hockey team to study the incidence of abnormal CMR findings suggestive of myocarditis in COVID-19 positive athletes as compared with athletes negative for COVID-19 infection.

Four members of the men’s hockey team joined the study team. The first meeting focused on the importance of partnership – given the clinical dilemma of whether to screen for myocarditis and the uncertainty of how to manage positive findings as well as potential concerns related to privacy, confidentiality and publication of findings. With the support of the hockey players (PK, KJ, TW, HW), the team’s coaches and leadership from Yale Athletic Medicine, we proceeded with a case-control study to assess myocarditis in hockey players recently infected with COVID-19 compared with lacrosse players who had never tested positive for COVID-19-either by history or twice weekly screening tests. Lacrosse players were selected as both sports are considered to be moderate static (II)-high dynamic (C) component sports and likely would have similar athletic adaptation of the heart. In anticipation of abnormal CMR findings of uncertain significance, the study team committed to a shared decision-making model, wherein all clinically relevant information would be reviewed by a team of cardiologists and athletic medicine specialists - in partnership with the player - to determine return-to-play should any asymptomatic athletes have findings suggestive of myocarditis.

All male hockey players known to be infected with COVID-19 and all male lacrosse players were contacted for participation via e-mail (Supplemental File 1). As per Yale Athletic Medicine clinical protocol, following recommendations from the literature, all hockey players had previously undergone evaluation with a 12-lead electrocardiogram (ECG), transthoracic echocardiogram (TTE), and troponin T level within 10-days following COVID-19 infection.^13^ As part of the study, all participants (both hockey and lacrosse players) underwent CMR imaging within a pre-specified 8 week timeframe from infection among cases. CMR protocols included cine imaging, pre and post contrast T1 mapping for extracellular volume (ECV) assessment, standard T2-weighted (STIR), T2-weighted mapping, T2*, and late gadolinium enhancement (LGE) sequences.^13^ (Supplemental File 2). Additionally, fast-strain encoded magnetic resonance (fast-SENC) imaging was obtained for assessment of global and segmental circumferential and longitudinal strain, which has been shown to detect subclinical left ventricular (LV) dysfunction.^14^ Subjects were assessed for presence of abnormality on T1-weighted (i.e. ECV or LGE) imaging and T2-weighted (i.e. standard T2-weighted or T2 mapping) imaging, per the updated 2018 Lake Louise Criteria.^13^ Additionally, we sent a brief anonymous follow-up survey to all participants to assess their comfort with the study and overall exerience (Supplemental File 3).

We describe the collaborative process with hockey players, the clinical characteristics and CMR findings between cases and controls. We used Fisher’s Exact test and unpaired T-tests for categorical and continuous variables respectively recognizing that given the small sample size, we were not powered to find a statistical difference. Statistical analysis was performed using GraphPad Prism version 8, San Diego, CA. We also report on survey feedback.

The Yale Institutional Review Board approved of the study. De-identified data available upon request.

## Results

The study team, inclusive of sports medicine internists, general cardiologists, an electrophysiologist, and experts in CMR, echocardiography and nuclear imaging, along with four hockey players, met three times over Zoom to establish the protocol and to discuss findings. Of the Yale hockey players infected with COVID, 13 expressed interest in the study and eight enrolled. Reasons for non-participation among the players included pre-existing cardiac condition (N=1) or inabilty to schedule CMR within the pre-specified timeline (N=4). Four men’s lacrosse players (controls) enrolled in the study.

Baseline characteristics were not significantly different between the two groups, with the exception of heart rate prior to CMR, which paradoxically was lower in the case cohort (Table 1). All but one of the cases had the same viral strain of COVID-19, with that variant being different by eight separate mutations. Any symptoms present among the cases were considered mild, and were determined to not warrant additional work-up beyond the ECG, echocardiogram and troponin evaluation already required by Yale University.

**Table 1.** Baseline Demographics

|  | Cases | Controls | p |
| --- | --- | --- | --- |
| Number | 8 | 4 |  |
| Age at CMR, years | 22.4 (21.7-23.0) | 21.9 (21.5-22.2) | 0.192 |
| Age at Dx, years | 22.3 (21.5-22.9) | N/A | N/A |
| Race: Non-white (%) | 0* | 0 | 1 |
| Time from Dx to CMR, days | 39 (37.8-41) | N/A | N/A |
| Male, % | 100 | 100 | 1 |
| Height, cm | 185.5 (178.8-188.6) | 182.5 (181.8-186.3) | 0.711 |
| Weight, kg | 83.1 (79.5-93.5) | 90.0 (87.0-92.0) | 0.515 |
| BSA, m<sup>2</sup> | 2.08 (1.99-2.19) | 2.15 (2.13-2.15) | 0.514 |
| Number of significant medical comorbidities | 0 (0-0) | 0 (0-0) | 1 |
| Number of cardiac medications | 0 (0-0) | 0 (0-0) | 1 |
| SBP | 121.5 (113.3-125.8) | 124 (120-130) | 0.488 |
| DBP | 64.5 (62-67) | 71 (67.3-74.8) | 0.117 |
| HR | 60 (54.8-67) | 78 (73.5-83.5) | 0.0376 |
| **COVID-19 information** |  |  |  |
| Duration of symptoms (days) | 5 (3.8-7) | N/A | N/A |
| Number of symptoms # | 2 (0-2.25) | N/A | N/A |
| Presence of Chest Pain/Heaviness, % | 50 | N/A | N/A |
| **Laboratory** |  |  |  |
| Troponin | <0.01 (<0.01-<0.01) | N/A | N/A |
| Creatinine | 1.0 (1.0-1.1) | 1.2 (1.0-1.2) | 0.506 |
| Hematocrit | 45 (44-47) | 46 (45-47) | 0.305 |

\*1 case subject had no race selected
#Symptoms assessed included: fever, myalgias, shortness of breath/cough, anaguesia, anosmia, chest pain

The median time to CMR after COVID-19 diagnosis was 39 days (interquartile range 38-41 days). Cases and controls had similar indexed LV end diastolic and end systolic volumes, whereas the end diastolic and systolic volumes were statistically greater in the case cohort. Indexed right ventricular (RV) end diastolic and end systolic volumes were significantly higher in the cases. There was no difference in LV and RV ejection fractions between the two groups. (Table 3). The majority of studies (87.5% of cases and 75% of controls) demonstrated a small focal area of mid-myocardial LGE at the inferior RV insertion site, comprising less than 3% of the myocardium in both groups. Only one of the cases demonstrated an additional small area of mid-myocardial to epicardial LGE at the basal inferolateral wall, which was interpreted as subtle and non-specific (Figure 1A). The global T1 mapping, ECV, and T2 values were normal in all cases. Additionally, there were no statistically significant differences in parameters for pre-contrast T1 mapping, ECV, LGE or T2-weighted imaging between the two groups. Overall, none of the subjects met imaging criteria for definite myocarditis by CMR per the updated Lake Louise Criteria^13^, which requires an abnormality in both a T1 and T2 weighted sequence.

**Table 2.** Baseline cardiac characteristics

|  | Cases | Controls | p-value |
| --- | --- | --- | --- |
| **ECG data\*** |  |  |  |
| HR (bpm) | 53 (47-60.3) | 60.5 (58-63) | 0.132 |
| PR interval (ms) | 164 (159-164) | 148 (146-161) | 0.990 |
| QTc interval (ms) | 406 (395-417) | 405 (401-411) | 0.660 |
| **Echo data** |  |  |  |
| LVEF by 3DE #, % | 57.5 (56.8-61) | N/A | N/A |
| Abnormal EF, % | 0 | N/A | N/A |
| GLS+, % | -20 (-20 - -18) | N/A | N/A |
| LVEDD, mm | 55.5 (52.3-56.3) | N/A | N/A |
| LVESD, mm | 36.5 (33.5-38) | N/A | N/A |
| LVEDV, mm<sup>3</sup> | 238 (180-257) | N/A | N/A |
| LVEDVi, mm<sup>3</sup>/m<sup>2</sup> | 114 (92-123) | N/A | N/A |
| LVESV, mm<sup>3</sup> | 83 (76-107) | N/A | N/A |
| LVESVi, mm<sup>3</sup>/m<sup>2</sup> | 41 (38-50) | N/A | N/A |
| LVSV, mL | 144 (103-167) | N/A | N/A |
| LVSVi, mL/m<sup>2</sup> | 71 (53-77) | N/A | N/A |
| LAVol, mL | 68 (52-77) | N/A | N/A |
| LAVoli, mL/m<sup>2</sup> | 30 (26-38) | N/A | N/A |

\* Note that controls only had ECG upon admission to school and did not have a repeat test prior

**Table 3.** CMR data

|  | Cases | Controls | p-value |
| --- | --- | --- | --- |
| Time between positive COVID-19 test and CMR | 39 (37.8-41) | N/A | N/A |
| LVEF, % | 56.5 (56-58.8) | 59 (56-62) | 0.477 |
| LVEDV, mL | 249 (217-270) | 208 (202-217) | 0.137 |
| LVEDVi, mL/m<sup>2</sup> | 118 (103-126) | 97 (94-102) | 0.0622 |
| LVESV, mL | 107 (87-117) | 85 (82-89) | 0.144 |
| LVESVi, mL/m<sup>2</sup> | 52 (41-56) | 40 (39-41) | 0.0733 |
| LVSV, mL | 147 (125-153) | 123 (114-135) | 0.235 |
| LVSVi, mL/m<sup>2</sup> | 67 (63-69) | 58 (53-64) | 0.142 |
| LVEDD, mm | 58 (54-60) | 50 (48-54) | 0.0367 |
| LVESD, mm | 40 (38-42) | 34 (30-37) | 0.0138 |
| Anteroseptal wall, mm | 8.5 (7-9) | 8.5 (8-9.8) | 0.354 |
| Posterolateral wall, mm | 9.5 (8.5-10) | 8 (8-9) | 1 |
| LV mass, g | 180 (150-182) | 169 (143-186) | 0.693 |
| LV mass index | 81 (75-86.3) | 78.5 (66.3-87.5) | 0.543 |
| Cardiac Output, L/min | 8.1 (7.2-8.6) | 7.5 (7.0-8.0) | 0.439 |
| Cardiac Index, L/min/m<sup>2</sup> | 3.6 (3.5-4.3) | 3.5 (3.2-3.8) | 0.375 |
| RVEDV, mL | 316 (299-330) | 262 (251-264) | 0.0418 |
| RVEDVi, mL/m<sup>2</sup> | 148 (142-163) | 122 (116-124) | 0.0298 |
| RVESV, mL | 160 (152-169) | 120 (112-129) | 0.0326 |
| RVESVi, mL/m<sup>2</sup> | 77.5 (69-84) | 56 (52-60) | 0.0254 |
| RVSV, mL | 153 (148-158) | 135 (126-142) | 0.159 |
| RVSVi, mL/m<sup>2</sup> | 71 (68.8-75.3) | 63 (58.3-67.3) | 0.110 |
| RVEF, % | 48.5 (47.8-51.3) | 51 (50-53.5) | 0.110 |
| LA size, mL | 25 (21-30.3) | 25.5 (24.8-26.8) | 0.888 |
| RA size, mL | 27.5 (25-29.5) | 27.5 (26.5-28) | 0.730 |
| **Parametric mapping** |  |  |  |
| % with papillar scar | 57.1 # | 75 | 1 |
| T1, pre-contrast, ms | 873 (861-889) | 874 (859-882) | 0.621 |
| T2 values, ms | 44.5 (44.1-45.3) # | 44.7 (44.3-45.1) | 0.956 |
| T2 ratios (heart/SM) | 1.58 (1.50-1.81) # | 1.63 (1.47-1.68) | 0.468 |
| T2*, ms | 38 (36.5-41) # | 38 (36.5-38) # | 0.583 |
| ECV, % | 24 (22.5-24) # | 23.5 (22.3-24.3) | 0.670 |
| **LGE segments** |  |  |  |
| 0 segments, % | 12.5 | 25 | 1 |
| 1+ segment, % | 87.5 | 75 | 1 |
| 2 segment, % | 12.5 | 0 | 1 |
| LV LGE % | 2.3 (2.2-3.1) | 2.6 (1.9-2.9) | 0.299 ^^ |
| Left Atrial LGE Volume (mm<sup>3</sup>) | 122 (83.5-204.5) | 29.5 (22.8-69.0) | 0.224 ^^ |
| **Strain** |  |  |  |
| **Left Atrial Strain (%)** |  |  |  |
| Systole | 45.9 (41.2-49.4) | 51.0 (47.6-53.2) | 0.319 |
| Passive Emptying | 16.6 (14.1-21.7) | 21.8 (17.5-25.8) | 0.272 |
| Mid-Ventricular diastole | 29.6 (24.1-31.4) | 27.4 (26.1-29.4) | 0.962 |
| **Left Atrial Strain rate (%/s)** |  |  |  |
| Systole | 1.9 (1.6-2.0) | 2.2 (1.9-2.5) | 0.0758 |
| Passive Emptying | -2.0 (-2.4- -1.9) | -2.5 (-2.5- -2.3) | 0.384 |
| Mid-Ventricular diastole | -1.5 (-1.8- -1.0) | -1.6 ( -2.0- -1.3) | 0.284 |
| **Fast-SENC strain+** |  |  |  |
| LV GLS, % | -15.3 (-15.5 - -13.9) | -16.0 (-17.5 - -15.2) | 0.161 |
| LV GCS, % | -17.5 (-19.1 - -17.4) | -17.4 (17.9 - -14.9) | 0.127 |
| RV GLS, % | -17.0 (-17.7 - -16.5) | -17.5 (-17.7 - -17.2) | 0.565 |
| RV GCS, % | -16.8 (-17.4 - -16.4) | -15.1 (-16.3 - -14.9) | 0.142 |

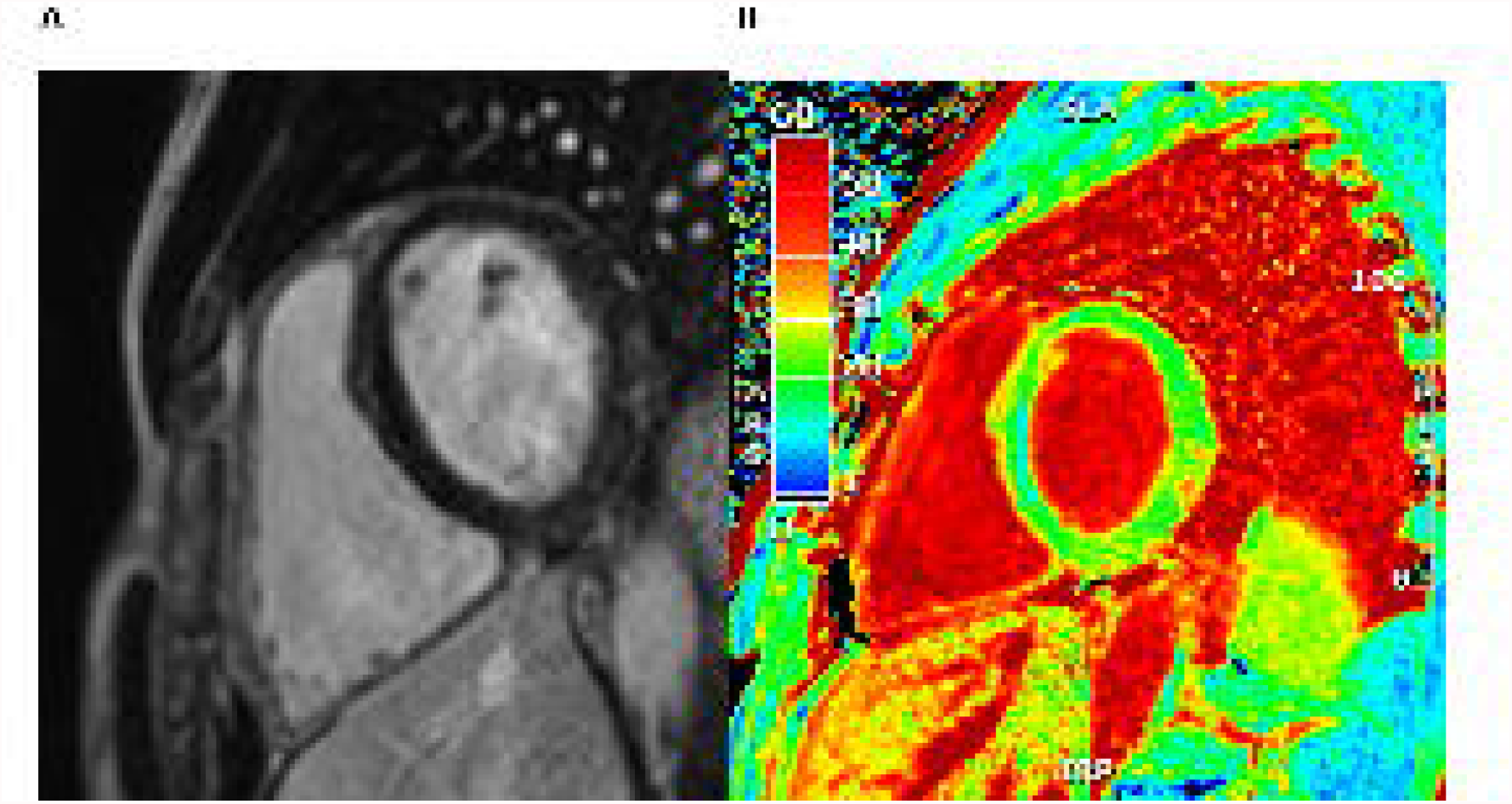

Additionally, there were no differences in left atrial LGE volume, left atrial strain, or papillary muscle fibrosis between the two groups. Left and right ventricular global longitudinal and global circumferential strain was similar between the two groups as well (Table 3), though two LV myocardial segments were significantly decreased in the cases and one segment of the RV was significantly increased in controls – a finding of unclear clinical significance (Supplemental Tables 1 and 2, Figure 1A).

Results were communicated back to all participants via telephone and MyChart messaging. No findings required meeting with the study physicians nor impacted return to play. For the athlete with late gadolinium enhancement outside of the RV insertion site, we communicated that without symptoms, the findings were non-specific and could not definitively be correlated with COVID-19 infection. We also communicated that while there are some data showing increased risk among “diseased” populations with LGE, we do not know if this data applies to functionally normal hearts, and that we had a low concern for any short or long-term sequelae. Overall, participants were appreciative of the information, based on informal feedback. All four controls and six of the eight case subjects participated in the follow-up survey. Nine of the ten respondents felt the study was important to learn about the effect of COVID-19 in athletes and five of the six cases responded that they felt comfortable/very comfortable participating knowing that their teammates and team physicians participated on the study team (Supplemental Figure).

## Discussion

In this study of collegiate athletes with asymptomatic or mildly symptomatic COVID-19, we sought to partner with athletes to understand the prevalence of CMR findings concerning for myocarditis, which could potentially have short and long-term effects, and to collaboratively develop protocols for communicating findings of uncertain significance with players. We found that athletes were understanding of the potential ambiguity of findings and of the tension between keeping players safe and allowing them to thrive in their athletic careers. Moreover, players were eager to be part of the protocols that could impact their health and return to play, desired transparency of information, and overall wanted to contribute to science. While prior studies have reported incidence of myocarditis in athletes with COVID-19, none have reported on how cases were clinically managed and how return to sports participation was decided.

Partnered research is important for establishing trust between clinicians, health systems and the community being studied.^15,16^ Citizen science – in which the general public (including patients) voluntarily and actively provides input and participates in the research – is increasinglyimplemented to rapidly advance research into COVID-19.^17^ This is in contrast to the traditional research process in which stakeholders typically do not have a say in shaping the questions or the research protocols to which they consent. By partnering with members of the hockey team in developing the protocol and procedures for relay of information, we allowed for active participation by the subjects and were able to address any concerns. For example, one player noted that even if the information was uncertain, it was best to share it with the player and report it in the medical record so they could use that information should any health concerns arise in the future, thus giving athletes the opportunity to make informed decisions regarding their health and career.

With respect to CMR findings, the predominant differences between cases and controls was in both left and right ventricular sizes, although we found no significant differences in the ejection fractions. Whether these differences are due to chance, athletic conditioning, or COVID-19 is unknown. Global strain values were similar between the two groups although some regional differences were seen. This could represent subclinical dysfunction, as fast-SENC has been shown to be predictive of future cardiac dysfunction, but findings are limited for interpretation given the small sample size and lack of long term clinical outcomes in this cohort. A small amount of LGE was observed in cases and controls, commonly mid-myocardial and at the RV insertion site. None of the subjects met definite imaging criteria for myocarditis by CMR, and there were no significant differences in tissue characterization between the cases and controls.

Our study does have significant limitations. Most notable is the small sample size.. Many athletes had left campus on or around IRB approval of the study due to the holiday and did not return in time to fall within the pre-specified eight week window. Additionally, we do not know the perspectives or concerns of athletes who chose not to participate, though the primary reasons given by non-participants in the study was a lack of time.

## Conclusions

Since the outset of the COVID-19 pandemic, several studies have demonstrated subclinical findings suggestive of myocarditis in asymptomatic or mildly symptomatic athletes with COVID-19. Partnering with study participants ahead of time and involving them in the study protocol assisted in participant autonomy and comfort level as well as management of research findings. Community outreach and participant involvement in study design should be considered for research involving athletes, and all other types of medical research.

## Data Availability

All data produced in the present study are available upon reasonable request to the authors.

## Acknowledgements

The authors would like to acknowledge the support of Francine LoRusso, RN, MHA and Keith Churchwell, MD.

Additionally, we would like to thank the Yale hockey and lacrosse teams for their participation in this study.

