## Supplemental File for "Partnering with Athletes to Assess Risk of COVID-Related Myocarditis"

Supplemental Table 1. Regional variation of left ventricular strain +

|  | Cases | Controls | p-value |
| --- | --- | --- | --- |
| **Longitudinal strain (%)** |  |  |  |
| *Basal Segments* |  |  |  |
| Anterior | -16.0 (-18.2 - -15.4) | -17.6 (-18.2 - -16.8) | 0.473 |
| Anteroseptal | -14.3 (-14.9 - -13.8) | -15.0 (-16.4 - -14.2) | 0.343 |
| Inferoseptal | -15.7 (-16.7 - -14.9) | -14.0 (-14.9 - -13.5) | 0.253 |
| Inferior | -18.6 (-19.4 - -16.9) | -15.5 (-17.6 - -14.6) | 0.305 |
| Inferolateral | -14.9 (-20.4 - -11.6) | -13.7 (-16.5 - -12.7) | 0.721 |
| Anterolateral | -16.5 (-18.5 - -15.4) | -16.8 (-17.1 - -15.2) | 0.588 |
| *Mid Segments* |  |  |  |
| Anterior | -14.9 (-15.3 - -12.8) | -13.0 (-16.7 - -12.9) | 0.481 |
| Anteroseptal | -14.7 (-15.3 - -14.2) | -18.0 (-18.9 - -16.6) | 0.163 |
| Inferoseptal | -14.2 (-16.0 - -13.0) | -19.1 (-19.8 - -17.4) | 0.0641 |
| **Inferior** | **-12.6 (-13.5 - -12.3)** | **-17.6 (-18.6 - -16.0)** | **0.00868** |
| Inferolateral | -14.3 (-15.9 - -11.5) | -18.2 (-18.3 - -16.6) | 0.0789 |
| Anterolateral | -15.0 (-15.6 - -14.1) | -13.5 (-17.5 - -13.2) | 0.632 |
| *Apical Segments* |  |  |  |
| **Anterior** | **-13.5 (-13.7 - -10.3)** | **-17.0 (-17.2 - -15.2)** | **0.0455** |
| Septal | -15.7 (-18.5 - -13.2) | -21.3 (-21.4 - -19.3) | 0.138 |
| Inferior | -15.0 (-17.4 - -13.9) | -16.5 (-18.1 - -14.6) | 0.638 |
| Lateral | -10.9 (-12.3 - -10.7) | -14.7 (-15.6 - -13.0) | 0.210 |
| **Circumferential strain (%)** |  |  |  |
| *Basal Segments* |  |  |  |
| Anterior | -13.5 (-15.4 - -12.6) | -13.7 (-13.9 - -13.3) | 0.826 |
| Anteroseptal | -15.5 (-16.3 - -14.4) | -184 (-19.1 - -16.1) | 0.234 |
| Inferoseptal | -15.3 (-16.6 - -14.6) | -16.2 (-16.6 - -15.2) | 0.946 |
| Inferior | -17.1 (-18.2 - -16.3) | -16.4 (-18.1 - -16.1) | 1 |
| Inferolateral | -20.1 (-20.8 - -17.5) | -17.6 (-19.0 - -17.6) | 0.590 |
| Anterolateral | -18.9 (-19.6 - -18.3) | -15.9 (-18.4 - -15.7) | 0.372 |
| *Mid Segments* |  |  |  |
| Anterior | -18.7 (-20.6 - -16.5) | -18.3 (-19.7 - -14.8) | 0.566 |
| Anteroseptal | -14.1 (-15.2 - -13.6) | -13.0 (-14.4 - -12.1) | 0.327 |
| Inferoseptal | -18.6 (-19.2 - -17.7) | -17.3 (-17.6 - -16.4) | 0.182 |
| Inferior | -19.6 (-20.3 - -17.4) | -14.1 (-16.9 - -11.7) | 0.102 |
| Inferolateral | -18.2 (-21.1 - -15.0) | -16.8 (-16.9 - -15.1) | 0.354 |
| Anterolateral | -21.1 (-22.2 - -20.8) | -19.3 (-20.9 - -15.7) | 0.204 |
| *Apical Segments* |  |  |  |
| Anterior (3Ch) | -18.3 (-20.2 - -16.9) | -16.2 (-17.2 - -14.0) | 0.169 |
| Anterior (2Ch) | -22.9 (-23.7 - -22.1) | -22.9 (-23.2 - -21.4) | 0.436 |
| Septal | -18.6 (-20.0 - -18.0) | -17.1 (-17.6 - -16.6) | 0.343 |
| Inferior | -19.3 (-20.8 - -15.8) | -9.9 (-14.2 - -8.8) | 0.0900 |
| Lateral (3Ch) | -18.1 (-20.2 - -14.3) | -10.5 (-14.3 - -8.2) | 0.184 |
| Lateral (4Ch) | -20.5 (-23.3 - -19.8) | -18.8 (-21.2 - -13.1) | 0.259 |
| Apical cap (3Ch) | -17.0 (-20.8 - -14.9) | -15.5 (-16.6 - -11.5) | 0.229 |
| Apical cap (4Ch) | -18.4 (-18.8 - -18.2) | -18.6 (-18.9 - -13.5) | 0.184 |
| Apical cap (2Ch) | -20.4 (-20.8 - -17.5) | -19 (-19.6 - -18.5) | 0.881 |

+ strain data was not available for one case and one control subject

3Ch: 3 chamber view, 2Ch: 2 chamber view, 4Ch: 4 chamber view

Supplemental Table 2. Regional variation of right ventricular strain+

|  | Cases | Controls | p-value |
| --- | --- | --- | --- |
| **Longitudinal strain (%)** |  |  |  |
| *Basal Segments* |  |  |  |
| Anterior | -16.2 (-18.9 - -15.8) | -17.3 (-17.6 - -16.6) | 0.898 |
| Lateral | -18.5 (-19.7 - -17.6) | -19.3 (-19.4 - -18.4) | 0.907 |
| Inferior | -17.1 (-17.7 - -16.5) | -19.1 (-19.5 - -18.9) | 0.258 |
| *Mid Segments* |  |  |  |
| Anterior | -16.2 (-17.1 - -15.8) | -15.7 (-16.0 - -15.7) | 0.574 |
| Lateral | -17.2 (-17.7 - -16.5) | -17.2 (-17.5 - -16.8) | 0.760 |
| Inferior | -15.8 (-16.2 - -14.7) | -16.8 (-17.2 - -16.4) | 0.113 |
| **Circumferential strain (%)** |  |  |  |
| *Basal Segments* |  |  |  |
| **Anterior** | **-17.1 (-17.6 - -16.0)** | **-13.3 (-14.9 - -13.0)** | **0.0390** |
| Lateral | -16.6 (-17.3 – 15.7) | -14.3 (-15.8 - -14.2) | 0.263 |
| *Mid Segments* |  |  |  |
| Anterior | -17.0 (-18.2 - -16.0) | -16.3 (-18.3 - -16.2) | 0.592 |
| Lateral | -18.2 (-19.6 - -15.9) | -16.8 (-17.6 - -15.0) | 0.342 |
| *Apical Segments* |  |  |  |
| Lateral | -16.1 (-17.4 - -15.9) | -16.5 (-16.7 - -15.0) | 0.354 |

+ strain data was not available for one case and one control subject

**Supplemental File 1.** Recruitment letter to hockey/lacrosse team captains, November 17, 2020.

Dear [Team Captain],

I am a cardiologist and part of the Yale Sports Cardiology Team. I am reaching out to see if you and your teammates would be interested in participating in a study of the cardiac effects of COVID-19.

As you may know, myocarditis (inflammation of the heart muscle) is one of the complications seen with COVID and some recent studies from Ohio State University (among athletes) and from Germany (general population) show that there are MRI abnormalities without symptoms of clinical myocarditis. This is of concern in competitive athletes since myocarditis can lead to dangerous heart rhythms and decreased heart function without rest. On the flip side, radiographic findings that are not supported by clinical symptoms are also potentially benign/incidental – and of no clinical consequence – and there is a risk that athletes may be subject to testing that is at best unnecessary, and at worst, detrimental to their well-being and success. Currently, there is no recommendation to do MRIs in all athletes with COVID, but there is still a lot of uncertainty. To study this question, we propose doing MRIs of the heart in the players who tested positive for COVID-19 compared with a control group of athletes who have not had COVID-19.

We are reaching out to see if you might be interested in being part of the study with a commitment to do one MRI in the next few weeks *and a second MRI of the heart ~3-12 months later.*

I am happy to talk more with the team or individual athletes who are interested.

Thanks so much for considering.

Best,

[Signature]

**Supplemental File 2.** Cardiac MRI imaging protocol.

CMR imaging occurred in the Yale New Haven Hospital 1.5 Tesla scanner (Aero, Avanto, Siemens Healthcare, Erlangen, Germany), with both standard clinical sequences and the strain research sequence performed within one scan, as outlined below. ECG gating was performed with electrode placement on the subject’s chest, and sequences will be breath-held, other than the 3D LGE, which will be respiratory navigator-gated.

Conventional Imaging Sequences: Cine imaging were acquired in the two, three, and four-chamber long axis cardiac views, along with contiguous short axis slices of the left ventricle from the base to apex. T2 weighted imaging and T2* imaging in the two, three, and four-chamber views as well as basal, mid, and apical short axis slices were obtained for assessment of inflammation and edema. T2 mapping with T2-weighted images of a higher spatial resolution than regular T2 weighted images were acquired in long axis and short axis views, and then a quantitative T2 map was automatically generated using an exponential fit.

2D Late Gadolinium Enhancement Imaging was then acquired in ventricular diastole, 8-10 min post gadolinium contrast administration, in the long axis views and in contiguous short axis slices of the left ventricle. 3D Late Gadolinium Enhancement Imaging of the whole heart was performed during mid ventricular diastole using an ECG-triggered and respiratory navigator-gated sequence, 15-20 minutes after administration of gadolinium contrast agent.

T1 mapping was performed in the two, three, and four-chamber views as well as basal, mid, and apical short axis slices before and 15 minutes after contrast injection, allowing for calculation of extracellular volume quantification of the myocardium.

Strain research sequence: Strain encoding (SENC) will be measured using the MyoStrain 5.1.1 software that was recently approved by the FDA. The images are obtained with short axis and long axis tagged images of the left ventricle, and a semi-automated post-processing system generated strain maps for assessment of myocardial deformation.

**Supplemental File 3**. Follow-up Survey

1. I am a:

a. Case subject (ie underwent MRI as a result of me having acquired COVID-19)
b. Control subject (ie at the point of my MRI, I had not been infected with COVID-19)

2. On a scale of 1 (strongly agree), 2 (somewhat agree), 3 (neutral), 4 (somewhat disagree), 5 (strongly disagree), please rate the degree to which you agree with the following statements:

a. This study was important for learning about the effects of COVID-19 in athletes.
b. I felt comfortable participating in the research study knowing that the results might provide more information about my health.
c. I felt nervous or worried about participating in the research study, knowing that the results might impact my return to training/playing.
d. I felt comfortably participating in the study, knowing my team members participated in the design of the study.
e. I felt comfortable participating in the study knowing my team doctors participated in the design of the study.
f. The results of my MRI were shared with me in a way that I could understand.
g. The results of my MRI influenced my confidence to return to training.

3. Were you concerned about the privacy of your information?

a. Very concerned
b. Somewhat concerned
c. Neutral
d. Not concerned

4. How did having athletes on the study team change your concern about the privacy of your data?

a. Increased my concern
b. Did not affect my concern
c. Decreased my concern
d. I was not concerned in the first place.

5. Please leave any other comments regarding this study, your experience or comfort level in participating for the study team.

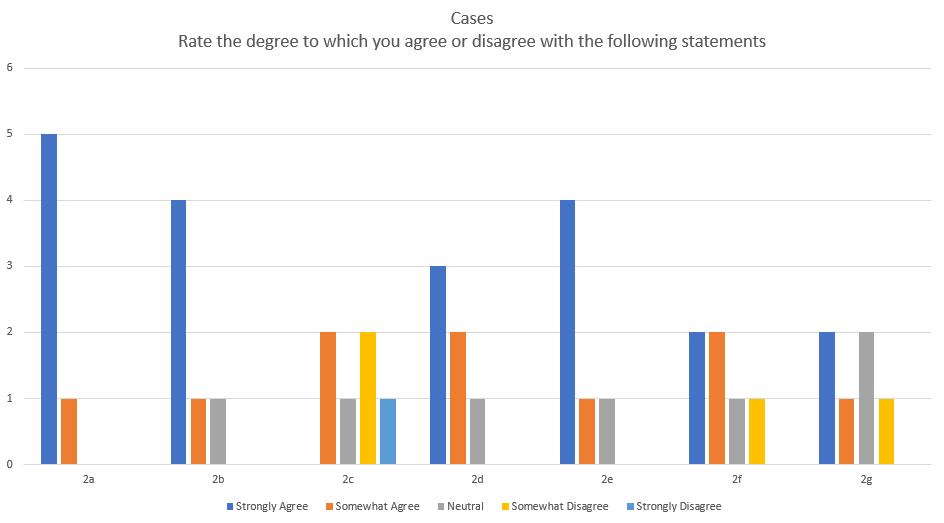

Supplemental Figure. Case responses to Question 2 of the survey.
